# Association between SARS-Cov-2 infection during pregnancy and adverse pregnancy outcomes: a re-analysis of data from Wei et al. (2021)

**DOI:** 10.1101/2021.06.22.21259302

**Authors:** Aho Glele Ludwig Serge, Emmanuel Simon, Camille Bouit, Maeva Serrand, Laurence Filipuzzi, Karine Astruc, Philippe Kadhel, Paul Sagot

## Abstract

**Background:** Wei et al. have published a meta-analysis (MA) which aimed to “evaluate the association between SARS-CoV-2 infection during pregnancy and adverse pregnancy outcomes”.

Using classical random-effects model, they found that SARS-CoV-2 infection was associated with preeclampsia, preterm birth and stillbirth.

Performing MA with low event rates or with few studies may be challenging as MA relies on several within and between study distributional assumptions.

**Methods:** to assess the robustness of the results provided by Wei et al., we performed a sensitivity analysis using several frequentist and Bayesian meta-analysis methods. We also estimated fragility indexes.

**Results:** For eclampsia (patients with Covid-19 vs without), the confidence intervals of most frequentist models contain 1. All beta-binomial models (Bayesian) lead to credible intervals containing 1. The prediction interval, based on DL method ranges from 0.75 to 2.38. The fragility index is 2 for the DL method.

For preterm, the confidence (credible) intervals exclude 1. The prediction interval is broad, ranging from 0.84 to 20.61. The fragility index ranges from 27 to 10.

For stillbirth, the confidence intervals of most frequentist models contain 1. Six Bayesian MA models lead to credible intervals containing 1.The prediction interval ranges from 0.52 to 8.49. The fragility index is 3.

**Interpretation:** Given the available data and the results of our broad sensitivity analysis, we can only suggest that SARS-CoV-2 infection during pregnancy is associated to preterm, and may be associated to preeclampsia. For stillbirth, more data are needed as none of the Bayesian analyses are conclusive.

## Background

Wei et al. (1) have recently published a systematic review and meta-analysis (MA) which aimed to “evaluate the association between SARS-Cov-2 infection during pregnancy and adverse pregnancy outcomes”.

The authors show that SARS-CoV-2 infection is associated with preeclampsia, preterm birth and stillbirth. They also stated that they used Mantel-Haenszel method, but the results of figure 2 are related to a random-effects inverse-variance model, with DerSimonian-Laird estimate of tau^2^ and continuity correction.

Higgin’s I2 was used to assess heterogeneity. Although this approach is widely mentioned, the point estimate I2 should be interpreted cautiously when a MA has few studies (2), and the confidence interval should be provided.

MA relies on several within and between study distributional assumptions that are sometimes hidden (3).

Several methods are available to assign weights in meta-analyses, e.g. Mantel-Haenszel for fixed-effect MA or inverse-variance for fixed-effect or random-effects MA. For random-effects MA, there are several ways to estimate the between-study variance, e.g. DerSimonian and Laird (DL), restricted maximum likelihood (REML). Finally, there are also different ways to estimate the confidence interval for the summary effect (e.g. Wald or Hartung-Knapp-Sidik-Jonkman (HKSJ) method). The same apply to confidence interval for the between-study variance (e.g. Q-Profile method). See (4,5) for more information.

Performing MA with low event rates or with few studies, or both, may be challenging. For example, estimating between-study heterogeneity is difficult in this case, and inaccurate estimation of this heterogeneity may lead to too narrow confidence intervals.

Statistical methods can never completely resolve the issue of sparse data. Different methods may give different results, and using a suboptimal approach may lead to erroneous conclusions (6).

## Methods

In order to assess the robustness of the results provided by Wei et al. (1), we performed a sensitivity analysis using a range of frequentist and Bayesian meta-analysis methods. We also estimated fragility indexes (and Fragility Quotient, which is fragility index divided by the total study sample size). This intuitive measure of the robustness of a trial, is a number indicating how many patients would be required to convert a trial from being statistically significant to not significant (7). This index has been adapted for MA (8) and is currently being refined methodologically (19). We estimated FI with DL method and restricted maximum likelihood with Knapp-Hartung (KNHA) test.

Wei et al. (1) reported three outcomes: preeclampsia, preterm and stillbirth. We focused on preeclampsia and stillbirth which contain sparse data.

For frequentist MA, several methods were used (9,10), including that of Hemni and Copas, which was found to be less sensitive to publication bias than DL method (11,12) (see tables). Although not optimal, continuity correction (0.5) was applied to studies with zero cells, in order to compare our results with the work of Wei et al. (1).

According to IntHout et al (18), we have also presented prediction interval, only for the model used by Wei et al. (1). It is the interval within the effect size of a new study would fall, if this study was selected at random from the same population of the studies already included in the MA.

For Bayesian MA, we used a binomial-normal hierarchical model (BNHM), i.e., modelling probabilities of success in each group (13), instead of modelling estimates of log odds-ratios directly (normal-normal model) for the between trial heterogeneity. We also used a beta-binomial (BB) model, which has shown good statistical properties for meta-analysis of sparse data (14,15). With these approaches, no continuity correction (of any type) is required as these models are based on the exact distributional assumptions unlike (commonly used) normal-normal hierarchical model. Several priors were used as sensitivity analysis (16,17).

Statistical analyses were performed with Stata (frequentist framework) and R software (Bayesian framework).

## Results

All estimates are shown in tables 1 and 2 and in appendix 1 (tables 3 and 4).

**Table 1:** sensitivity analysis performed using different methods to estimate the association between Covid-19 and preeclampsia

| Model/method | Odds ratio (95 % CI) | I2 (95 % CI) | Tau2 (95 % CI) |
| --- | --- | --- | --- |
| Patients with Covid-19 vs without Covid-19 |  |  |  |
| Frequentist |  |  |  |
| DL | 1.33 (1.03 to 1.73) | 30.9% (0.0 to 64.3) | 0.05 |
| DL-pred | 1.33 (0.75 to 2.38) |  |  |
| Perm | 1.33 (0.95 to 1.66) | 30.9% | 0.05 |
| Boot | 1.34 (1.02 to 1.78) | 31.0% (0.0 to 64.5) | 0.06 |
| REML | 1.36 (1.00 to 1.85) | 30.9% (0.0 to 64.3) | 0.09 (0.00 to 0.93) |
| MP | 1.35 (1.01 to 1.80) | 30.9% (0.0 to 64.3) | 0.07 (0.00 to 0.87) |
| SJ | 1.44 (0.94 to 2.20) | 30.9% (0.0 to 64.3) | 0.28 (0.00 to 0.87) |
| RO | 1.03 (0.98 to 1.81) | 30.9% | 0.05 |
| HKSJ | 1.33 (0.98 to 1.81) | 30.9% | 0.05 |
| KR (oim) | 1.36 (0.75 to 2.45) | 44.5% (0.0 to 85.5) | 0.09 (0.00 to 0.67) |
| KR (eim) | 1.36 (0.81 to 2.29) | 44.5% (0.0 to 85.5) | 0.09 (0.00 to 0.67) |
| PL-B | 1.33 (0.84 to 2.54) | 0.0% (0.0 to 75.9) | 0.0 (0.00 to 0.36) |
| PL-S | 1.33 (0.86 to 2.52) | 0.0% (0.0 to 75.9) | 0.0 (0.00 to 0.36) |
| HC | 1.33 (0.45 to 3.90) | 30.9% (0.0 to 64.3) | 0.05 |
| IVHet | 1.33 (0.98 to 1.99) | 30.9% | 0.05 |
| Sens (60) | 1.40 (0.97 to 2.02) | 60% 2(6.47 to 78.24) | 0.17 |
| Sens (75) | 1.46 (0.92 to 2.30) | 75% (75 to 85.5) | 0.34 |
| Bayesian |  |  |  |
| BNHM (WIP) | 1.40 (1.02 to 2.25) |  |  |
| BB (half normal) | 1.62 (0.88 to 3.00) |  |  |
| BB (uniform) | 1.60 (0.89 to 2.94) |  |  |
| BB (half Cauchy) | 1.60 (0.88 to 2.97) |  |  |
| BNHM (Vag) | 1.40 (1.02 to 2.18) |  |  |
| BN (t) | 1.47 (1.00 to 2.50) |  |  |
| Patients with severe vs mild Covid-19 |  |  |  |
| Frequentist |  |  |  |
| DL | 4.16 (1.55 to 11.15) |  |  |
| DL-pred | 4.16 (0.84 to 20.61) |  |  |
| Perm | Not computed: number of studies is below 6 |  |  |
| Boot | 4.16 (1.55 to 11.16) | 0.08% (0.0 to 79.2) | 0.001 |
| REML | 4.16 (1.55 to 11.15) | 0% (0.0 to 77.2) | 0 (0 to 4.65) |
| MP | 4.16 (1.55 to 11.15) | 0% (0.0 to 74)* | 0 (0 to 3.90) |
| SJ | 4.16 (1.55 to 11.15) | 0% | 0 |
| RO | 4.16 (1.11 to 15.59) | 0% | 0 |
| HKSJ | 4.16 (1.03 to 16.80) | 0% | 0 |
| KR (oim) | 4.16 (0.69 to 25.0) | 0% (0.0 to 77.2) | 0 (0 to 4.65) |
| KR (eim) | 4.16 (0.08 to 226.82) | 0% (0.0 to 77.2) | 0 (0 to 4.65) |
| PL-B | 4.16 | 0% (0.0 to 66.9) | 0 (0 to 2.78) |
| PL-S | 4.16 (1.23 to 27.24) | 0% (0.0 to 66.9) | 0 (0 to 2.78) |
| HC | 4.16 (1.66 to 10.40) | 0% | 0 |
| IVHet | 4.16 (1.55 to 11.15) |  |  |
| Sens(60) | 5.30 (1.01 to 27.80) | 60% (0.00 to 85.04) | 2.06 |
| Sens(75) | 5.60 (0.692 to 45.38) | 75% (38.36 to 89.86) | 4.12 |
| Bayesian |  |  |  |
| BNHM (WIP) | 4.48 (1.51 to 13.46) |  |  |
| BB (half normal) | 3.38 (0.92 to 13.06) |  |  |
| BB (uniform) | 3.29 (0.91 to 12.55) |  |  |
| BB (half Cauchy) | 3.38 (0.91 to 13.46) |  |  |
| BNHM (Vag) | 4.71 (1.55 to 14.44) |  |  |
| BN(t) | 8.10 (0.65 to 29.04) |  |  |

CI: confidence interval (frequentist) or credible (bayesian). I2: Higgin's I2; Tau2: between-study variance.
Boot: Bootstrapped DL; DL: DerSimonian-Laird); DL-pred: prediction interval with DL; HKSJ: Hartung-Knapp-Sidik-Jonkman; HC: Henmi and Copas; IVHet: Inverse-variance heterogeneity; KR: Kenward-Roger; MP: Mandel-Paule; Perm: permutation; PL: Partial Likelihood (-B: with Bartlett's correction; -S: with Skovgaard's correction); REML: Restricted Maximum Likelihood; RO: DL with SJ robust variance estimator; SJ: Sidik-Jonkman; Sens: sensitivity analysis (percentage of I2).
BB: Beta-Binomial; BNHM: Binomial-Normal Hierarchical Model; Vag: Vague prior; BN(t): Binomial Normal (t distribution); WIP: Weakly Informative Prior.

**Table 2:** sensitivity analysis performed using different methods to estimate the association between Covid-19 and stillbirth

| Model/method | Odds ratio (95 % CI) | I2 (95 % CI) | Tau2 (95 % CI) |
| --- | --- | --- | --- |
| Patients with Covid-19 vs without Covid-19 |  |  |  |
| Frequentist |  |  |  |
| DL | 2.11 (1.14 to 3.90) | 24.02% (0.00 to 67.77) | 0.15 |
| DL-pred | 2.11 (0.52 to 8.49) |  |  |
| Perm | 2.10 (0.98 to 4.24) | 24.02% (0.00 to 67.77) | 0.15 |
| Boot | 2.16 (1.08 to 4.32) | 28.94% (0.00 to 70.86) | 0.24 |
| REML | 2.07 (1.16 to 3.67) | 19.5% (0.00 to 87.2) | 0.12 (0.00 to 3.30) |
| MP | 2.12 (1.12 to 4.0) | 26% (0.00 to 93.5) | 0.17 (0.05 to 6.94) |
| SJ | 2.11 (1.12 to 3.96) | 25.2% | 0.16 |
| RO | 2.10 (0.99 to 4.46) | 24.0% | 0.15 |
| HKSJ | 2.10 (0.93 to 4.77) | 24.0% | 0.15 |
| KR (oim) | 2.07 (0.00 to 9.3e+27) | 19.5% (0.00 to 87.2) | 0.12 (0.00 to 3.30) |
| KR (eim) | 2.07 (0.00 to ?) | 19.5% (0.00 to 87.2) | 0.12 (0.00 to 3.30) |
| PL-B | 1.80(0.57 to 8.18) | 0.0% (0.0 to 71.3) | 0.00 (0.00 to 1.20) |
| PL-S | 1.80(0.61 to 6.74) | 0.0% (0.0 to 71.3) | 0.00 (0.00 to 1.20) |
| HC | 1.80 (0.52 to 6.29) | 24.0% | 0.15 |
| IVHet | 1.80 (0.87 to 3.75) | 24.0% | 0.15 |
| Sens (60) | 2.21 (0.85 to 5.76) | 60.0% (1.82 to 83.70) | 0.73 |
| Sens (75) | 2.18 (0.65 to 7.24) | 75.0% (43.39 to 88.96) | 1.45 |
| Bayesian |  |  |  |
| BNHM (WIP) | 1.84 (0.92 to 3.63) |  |  |
| BB (half normal) | 1.39 (0.46 to 3.74) |  |  |
| BB (uniform) | 1.40 (0.47 to 3.86) |  |  |
| BB (half Cauchy) | 1.39 (0.46 to 3.82) |  |  |
| BNHM (Vag) | 1.86 (0.98 to 3.60) |  |  |
| BN (Vag) | 1.96 (0.38 to 5.24) |  |  |

CI: confidence interval (frequentist) or interval (bayesian). I2: Higgin's I2; Tau2: between-study variance.
Boot: Bootstrapped DL; DL: DerSimonian-Laird); DL-pred: prediction interval with DL; HKSJ: Hartung-Knapp-Sidik-Jonkman; HC: Henmi and Copas; IVHet: Inverse-variance heterogeneity; KR: Kenward-Roger; MP: Mandel-Paule; Perm: permutation; PL: Partial Likelihood (-B: with Bartlett's correction; -S: with Skovgaard's correction); REML: Restricted Maximum Likelihood; RO: DL with SJ robust variance estimator; SJ: Sidik-Jonkman; Sens: sensitivity analysis (pourcentage of I2).
BB: Beta-Binomial; BNHM: Binomial-Normal Hierarchical Model; VAG: Vague prior; BN(t): Binomial Normal (t distribution); WIP: Weakly Informative Prior.

In some cases, no estimation was possible: this is the case for permutations, when less than six studies were included in the meta-analysis. Under these conditions, Bayesian estimates are more appropriate.

For eclampsia in general (comparing patients with Covid-19 vs without Covid-19), the confidence intervals of most frequentist MA models contain 1 (table 1).

The prediction interval, based on DL method ranges from 0.75 to 2.38.

The fragility index is 2 for the DL method used by the authors and the same with the REML-HKSJ method.

As for the Bayesian approach, all three beta-binomial models of MA lead to credible intervals containing 1. For binomial normal models, credible intervals do not contain one, but the upper bound of the interval is very close to 1.

For eclampsia when comparing patients with severe vs mild Covid-19, the frequentist sensitivity analysis confirms the authors’ results, except for one model out of 12 (Kenward-Roger’s model).

The prediction interval, based on DL method ranges from 0.84 to 20.61.

The fragility index is 2 for DL method and 1 with REML-HKSJ method.

When an heterogeneity of 75% is taken into account, the difference between the two groups is no longer significant.

Concerning the Bayesian analysis, only the binomial-normal models confirmed Wei et al. (1) initial results. None of the three beta binomial models results in credible intervals excluding 1.

For preterm (global analysis comparing patients with Covid-19 vs without Covid-19, or subgroup analysis comparing patients with symptomatic vs asymptomatic Covid-19), all frequentist models used in our sensitivity analysis lead to confidence or credible intervals excluding 1, except inverse variance heterogeneity model and Hemni-Copas’s method (table 3, appendix).

But the prediction interval, based on DL method is very broad, ranges from 0.84 to 20.61.

The fragility index ranges from 27 (global analysis with for the DL method) to 10 (subgroup analysis comparing symptomatic vs asymptomatic patients, with REML-HKNHA method). Details on these fragility indexes and quotients are in table 4 (Appendix).

All Bayesian credible intervals also exclude 1.

Finally for stillbirth, the confidence intervals of most frequentist meta-analysis (MA) models contain 1 (table 2).

The prediction interval, based on DL method ranges from 0.52 to 8.49.

The fragility index is 3 for the DL method and 3 with REML-HKNHA method.

As for the Bayesian approach, all six MA models lead to credible intervals containing one.

## Interpretation

We performed a large sensitivity study taking into account different approaches (frequentist and Bayesian) and different statistical models in order to estimate several parameters including odds ratio, I2, Tau and their respective confidence intervals.

With the exception of preterm infants, regardless of the frequentist or Bayesian statistical approach, confidence intervals of odds ratio frequently overlap 1 and thus show no association between Covid-19 and the occurrence of eclampsia or stillbirth. The prediction intervals are very wide and contain 1, for all three endpoints (eclampsia, stillbirth and preterm). Finally, the fragility indexes are only 1, 2 or 3 for eclampsia and stillbirth.

Concerning our Bayesian sensitivity analysis, our results are robust to variation of the priors.

Some authors have proposed the use of generalized linear mixed model (GLMM) to perform MA with sparse data (20). We have not used GLMM. Here again, several approaches are available. We considered that our sensitivity analysis was sufficiently broad. Finally, we did not take into account the quality of the studies via risk-of-bias analysis, which is possible through meta-regression.

## Conclusion

Given the available data and the results of our broad sensitivity analysis we can only suggest that SARS-CoV-2 infection during pregnancy is associated to preterm, and may be associated to preeclampsia. For stillbirth, more data are needed as none of the Bayesian analyses are conclusive.

## Data Availability

Data will be found in the original article by Wei et al.

## Contributions

Ludwig Serge Aho Glele (LSAG) and Emmanuel Simon (EM) contributed to the conception and design of the study.

LSAG performed the statistical analysis.

LSAG, EM,Philippe Kadhel and Paul Sagot drafted the manuscript.

Camille Bouit, Maeva Serrand, Laurence Filipuzzi and Karine Astruc reviewed the manuscript.

All of the authors contributed to the interpretation of the data, gave final approval of the version to be published and agreed to be accountable for all aspects of the work.

---

**Appendix, Table 3:**
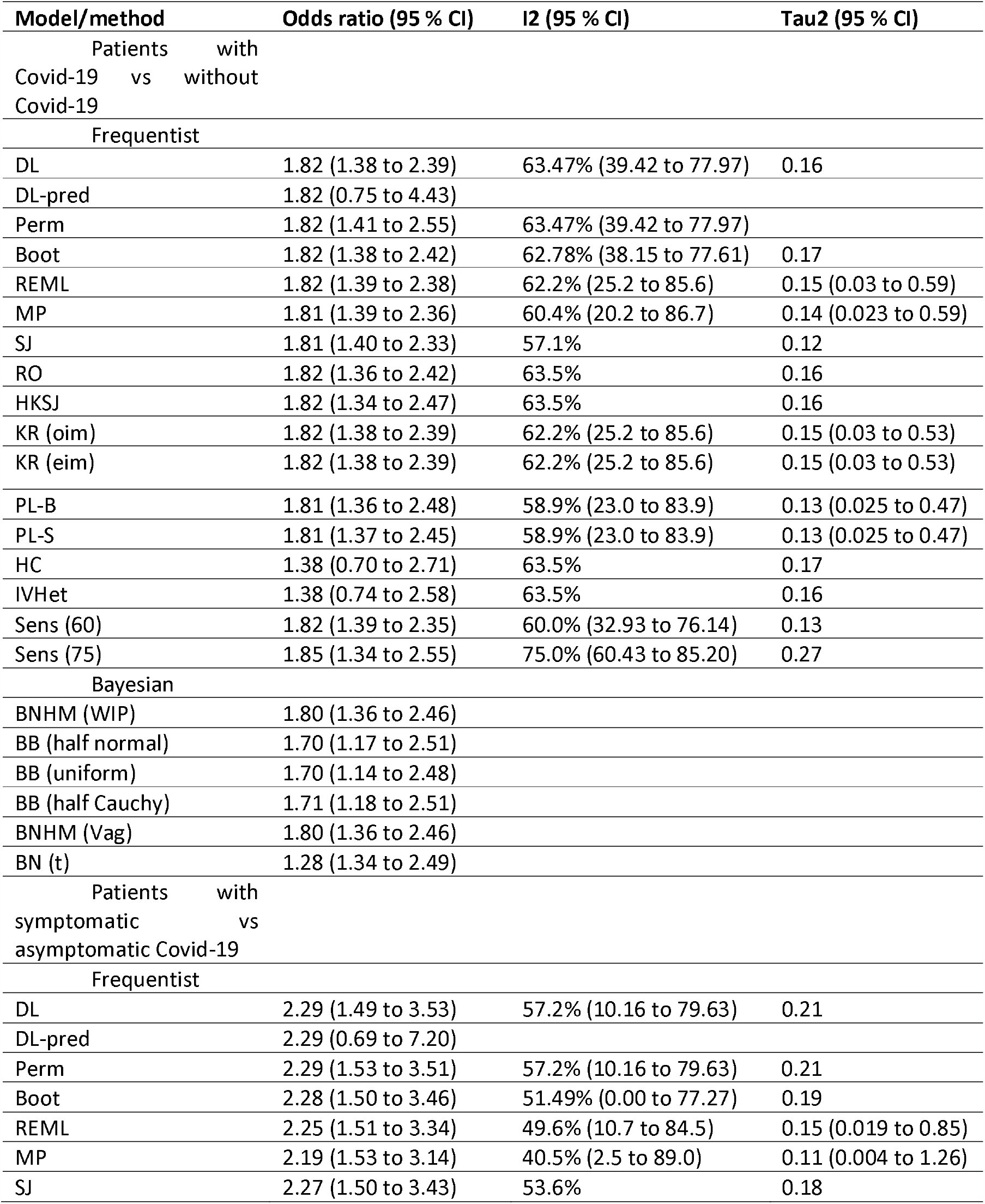
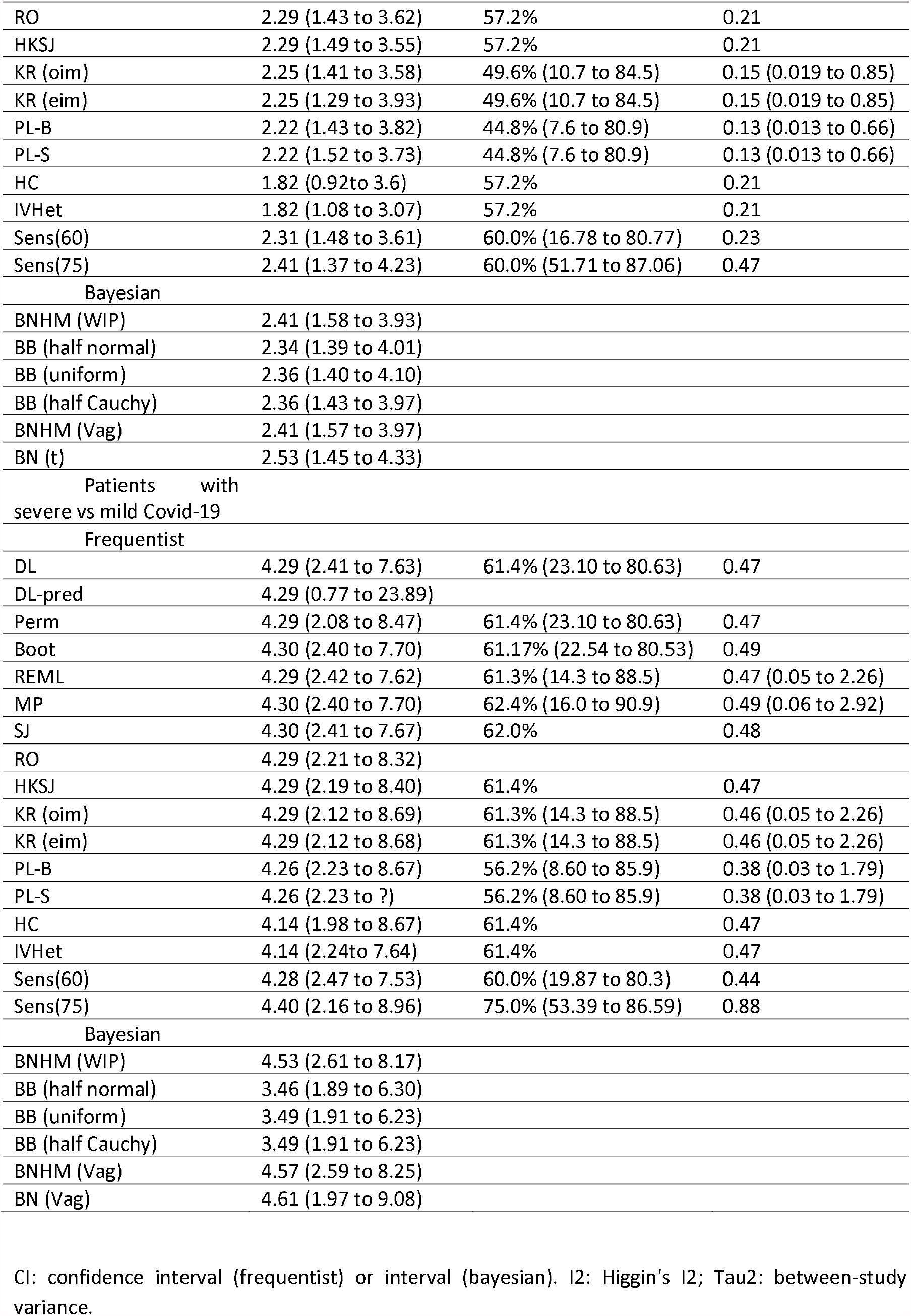
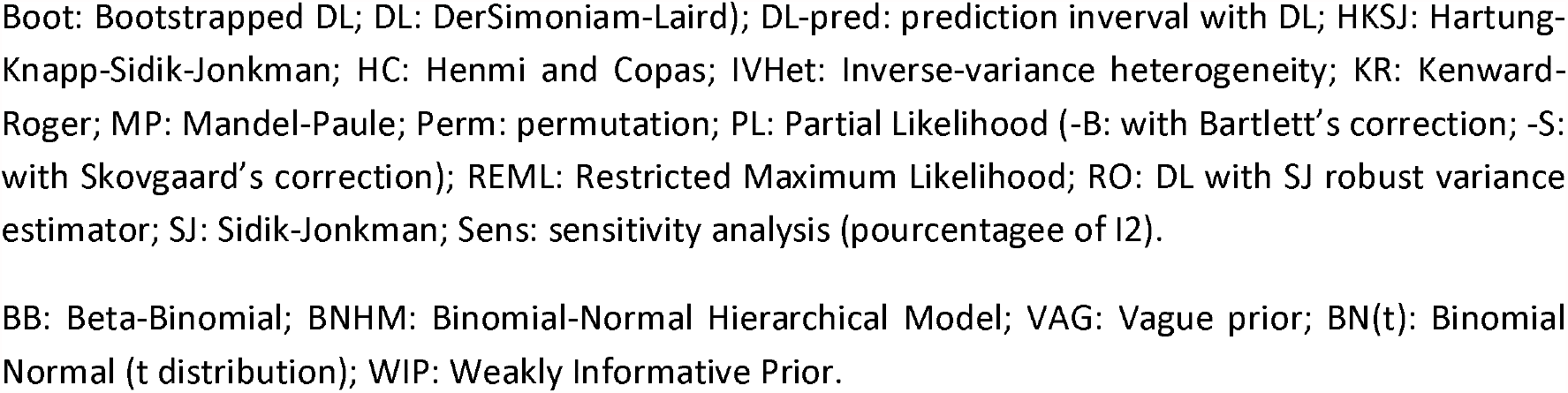
sensitivity analysis performed using different methods to estimate the association between Covid-19 and preterm birth

**Appendix, Table 4:** fragility index and fragility quotient

| **Diseases** | **Statistical method** | **Fragility index** | **Fragility quotient (%)** |
| --- | --- | --- | --- |
| **Eclampsia** |  |  |  |
| Covid-19 vs without Covid-19 | DL | 2 | 0 |
|  | REML-KNHA | 2 | 0 |
| Severe vs mild Covid | DL | 2 | 0.4 |
|  | REML-KNHA | 1 | 0.2 |
| **Stillbirth** |  |  |  |
|  | DL | 3 | 0.0 |
|  | REML-KNHA | 1 | 0.0 |
| **Preterm** |  |  |  |
| Covid-19 vs without Covid-19 | DL | 27 | 0.0 |
|  | REML-KNHA | 18 | 0.0 |
| Symptomatic vs asymptomatic Covid-19 | DL | 17 | 0.4 |
|  | REML-KNHA | 10 | 0.2 |
| Severe vs mild Covid-19 | DL | 20 | 1.4 |
|  | REML-KNHA | 13 | 0.9 |

DL: Dersimonian-Laird; REML: Restricted Maximum Likelihood (KNHA: Knapp- Hartung).

## References

1. Wei SQ, Bilodeau-Bertrand M, Liu S, Auger N. The impact of COVID-19 on pregnancy outcomes: a systematic review and meta-analysis. CMAJ [Internet]. 2021 Jan 1 [cited 2021 Mar 25]; Available from: https://www.cmaj.ca/content/early/2021/03/18/cmaj.202604

2. von Hippel PT. The heterogeneity statistic I2 can be biased in small meta-analyses. BMC Med Res Methodol [Internet]. 2015 Apr 14 [cited 2019 Mar 11];15. Available from: https://www.ncbi.nlm.nih.gov/pmc/articles/PMC4410499/

3. Jackson D, White IR. When should meta-analysis avoid making hidden normality assumptions? Biometrical Journal Biometrische Zeitschrift. 2018 Nov;60(6):1040.

4. Deeks JJ, Higgins JPT, Altman DG. Analysing data and undertaking meta-analyses. In: Higgins JPT, Thomas J, Chandler J, Cumpston M, Li T, Page MJ, et al., editors. Cochrane Handbook for Systematic Reviews of Interventions [Internet]. 2nd ed. Hoboken, NJ: Wiley-Blackwell; 2019. p. 241-284 (chapter 10). Available from: doi:10.1002/9781119536604

5. Veroniki AA, Jackson D, Viechtbauer W, Bender R, Bowden J, Knapp G, et al. Methods to estimate the between-study variance and its uncertainty in meta-analysis. Res Synth Methods. 2016 Mar;7(1):55–79.

6. Efthimiou O. Practical guide to the meta-analysis of rare events. Evidence-Based Mental Health. 2018 May 1;21(2):72–6.

7. Walsh M, Srinathan SK, McAuley DF, Mrkobrada M, Levine O, Ribic C, et al. The statistical significance of randomized controlled trial results is frequently fragile: a case for a Fragility Index. Journal of Clinical Epidemiology. 2014 Jun;67(6):622–8.

8. Atal I, Porcher R, Boutron I, Ravaud P. The statistical significance of meta-analyses is frequently fragile: definition of a fragility index for meta-analyses. J Clin Epidemiol. 2019 Jul;111:32–40.

9. Seide SE, Röver C, Friede T. Likelihood-based random-effects meta-analysis with few studies: empirical and simulation studies. BMC Medical Research Methodology. 2019 Jan 11;19(1):16.

10. Veroniki AA, Jackson D, Bender R, Kuss O, Langan D, Higgins JPT, et al. Methods to calculate uncertainty in the estimated overall effect size from a random-effects meta-analysis. Res Synth Methods. 2019 Mar;10(1):23–43.

11. Henmi M, Hattori S, Friede T. A confidence interval robust to publication bias for random-effects meta-analysis of few studies. arXiv:200207598 [stat] [Internet]. 2020 Jul 11 [cited 2021 Apr 12]; Available from: http://arxiv.org/abs/2002.07598

12. Henmi M, Copas JB. Confidence intervals for random effects meta-analysis and robustness to publication bias. Statistics in Medicine. 2010;29(29):2969–83.

13. Hamza TH, van Houwelingen HC, Stijnen T. The binomial distribution of meta-analysis was preferred to model within-study variability. J Clin Epidemiol. 2008 Jan;61(1):41–51.

14. Mathes T, Kuss O. A comparison of methods for meta-analysis of a small number of studies with binary outcomes. Res Synth Methods. 2018 Sep;9(3):366–81.

15. Mathes T, Kuss O. Beta-binomial models for meta-analysis with binary outcomes: Variations, extensions, and additional insights from econometrics. Research Methods in Medicine & Health Sciences. 2021 Mar 1;2(2):82–9.

16. Günhan BK, Röver C, Friede T. Random-effects meta-analysis of few studies involving rare events. Res Synth Methods. 2020 Jan;11(1):74–90.

17. Röver C, Bender R, Dias S, Schmid CH, Schmidli H, Sturtz S, et al. On weakly informative prior distributions for the heterogeneity parameter in Bayesian random-effects meta-analysis. Res Syn Meth. 2021 Feb 15;jrsm.1475.

18. IntHout J, Ioannidis JPA, Rovers MM, Goeman JJ. Plea for routinely presenting prediction intervals in meta-analysis. BMJ Open. 2016 Jul 1;6(7):e010247.

19. Lin L. Factors that impact fragility index and their visualizations. J Eval Clin Pract. 2021 Apr;27(2):356–64.

20. Ju K, Lin L, Chu H, Cheng L-L, Xu C. Laplace approximation, penalized quasi-likelihood, and adaptive Gauss-Hermite quadrature for generalized linear mixed models: towards meta-analysis of binary outcome with sparse data. BMC Med Res Methodol. 2020 Jun 11;20(1):152.

